# Early activity and impact of a neighbourhood multidisciplinary team that integrates health, and social support for underserved children and young people in Birmingham, UK: an observational study

**DOI:** 10.1101/2025.07.31.25332535

**Authors:** C Bird, F Dutton, S Kaur, C Wolhuter, I Litchfield

**Affiliations:** Birmingham Women’s and Children’s NHS Foundation Trust; Nuffield Department of Primary Care Health Sciences, University of Oxford; Small Heath Medical Practice, Birmingham, UK; GreenSquareAccord, UK; Department of Applied Health Sciences, University of Birmingham, UK; Birmingham Health Partners, Birmingham, UK

**Keywords:** Child health, Health policy, Health services research, Adolescent health, Social work

## Abstract

**Objectives:** The Sparkbrook Children’s Zone (SCZ) is an integrated model of care for children and young people in an economically marginalised area of Birmingham, United Kingdom. It is an early example of the National Health Service’s planned shift toward Neighbourhood Multi-Disciplinary Teams. This work presents early data on the uptake and impact of the three key components of the SCZ’s care offer: Preventative care, Clinical care, and Social support to help inform commissioners, social, and healthcare staff developing similar models.

**Methods:** Descriptive, observational study using routinely collected data to evaluate the activity and impact of SCZ weekly clinics (<16 years) in the three key care components.

**Results:** Demographics: From March 2022-December 2024, 2,265 CYP were booked into clinics (93.5% slots taken up); 45% <5 years; 89% of families from bottom IMD quintile. *Preventative health care:* Immunisation advice increased over the course of the opening months from 10.7% in 2023 to 40.2% in 2024 (compared to 1% in the surrounding primary care network); oral health promotion from 29.2% to 46.8%; smoking cessation advice from 1.8%% to 12.5%; 97% eligible children received Healthy Start vitamins; 70% had Body Mass Index measured. The Healthier Together health information app was delivered via text message to 100% of families after clinic visit. *Clinical care:* 73.8% of CYP discharged after clinical consultation, 3.8% referred to secondary care. 14.7% patients were not brought to appointments; *Social support:* 28.2%) CYP referred to family support worker, with the five most frequent reasons for referral being feeding issues, behaviour, activities, Special Educational Needs and Disabilities, and parenting skills.

**Conclusion:** Although preliminary, this initial data has offers valuable insight into patterns of disposition and referral including the increase in health promotion in a neighbourhood health offer. It has also raised awareness of the data set needed for long-term evaluation.

## Introduction

Children, young people (CYP) and their families living in high income countries face mounting challenges to their health and well-being, as the prevalence of chronic conditions, obesity, and mental ill health continues to increase [1]. These challenges are exacerbated in minoritized and economically-deprived communities by a range of socio-economic and cultural pressures that inhibit access and utilization of primary or preventative health care services [2, 3]. This has led to a rise in children’s attendances to emergency departments, frequently due to conditions that could be more effectively treated in community settings [4, 5].

In order to more effectively manage the health needs of CYP in underserved communities and reduce inappropriate ED attendance, a number of models of culturally sensitive and integrated care have emerged in North America, Europe, and Australia [6]. They share the broad aim of providing widely accessible health and social care or support for CYP that also mitigates the social determinants of ill-health such as poor housing, domestic violence, or food poverty [6, 7]. It is crucial that the design and delivery of these complex interventions are evidence-based yet the accrual of evidence in support of their design has been hindered by fragmented data sets, imprecision over key indicators, and the tension between aims of long-term improvement and the need to demonstrate short-term gains [8]. The result is that evidence of the benefits of integrated care remains inconsistent, particularly amongst underserved CYP [6, 9].

In the United Kingdom (UK) the potential of localised integrated health and social care has been recognised in the NHS Long Term Plan which has seen the recent introduction of the national framework for integrated neighbourhood teams for CYP and in the UK government’s 10-year plan for health [10]. However, UK efforts at integrating health and social care to date have tended to focus on high-level processes such as integrated commissioning, strategic planning and there is a lack of guidance from the NHSE on how best to design and deliver place based integrated care services along with practical support [11, 12]. This includes which combinations of medical and social care/support services are most effective at reaching underserved CYP; how to merge hard-pressed care organisations with differing agendas and funding mechanisms; and the practical support from a service perspective necessary to ensure delivery.

One NHS-funded pilot offers a useful early example of integrated neighbourhood care delivered by local health and social care providers for CYP in an underserved population in Birmingham [11, 13]. Called the “Sparkbrook Children’s Zone” the service has been used as exemplar of integrated neighbourhood health in a 2025 report by NHS England [10]. To help support the development of similar initiatives the work presented here describes the details of the service offering and explores service utilisation from the first 34 months since it opened in March 2022 to understand how effectively the service is delivering its 3 key components preventative care, clinical care, and social support for CYP.

## Methods

### Study design

An observational study, presenting early quantitative data of delivery and take-up of the SCZ service. The work presented here forms part of a mixed methods evaluation funded by Birmingham Women’s and Children’s NHS foundation Trust and led by the Department of Applied Health Sciences, University of Birmingham [11, 13]. The work was supported by the members of our public patient involvement panel who provided advice and feedback throughout the design and reporting of this study.

### Setting/study population

Birmingham is a large, diverse city in the West Midlands, UK, with a population of over 1.1 million. The Sparkbrook and East Balsall Heath district is the second most populous ward with a young population (35% are <18 years compared to 20% nationally), high diversity and one of the highest rates of infant mortality, at 8/1,000 deaths compared to 3.3/1,000 nationally [14].

The ward was selected for a pilot integrated care service for CYP, based on a high level of emergency department attendances from the district and high level of deprivation, meeting the NHS’s priorities around tackling health inequalities [15]. It was funded by NHS England’s Integrated Models of Care and Early Years programmes, with Birmingham Women’s and Children’s NHS Foundation Trust acting as the lead organisation for the pilot.

### Sparkbrook Children’s Zone

The service is available to CYP <16 years old and led by three key organisations: GreenSquareAccord, a national housing and social care provider which is currently the Voluntary Care Sector (VCS) Early Help Lead in the locality (for CYP 0-19 years) [16]; health care professionals from Birmingham Women’s and Children’s NHS Foundation Trust; and GPs drawn from the local primary care network (Balsall Heath, Spark Hill and Moseley Primary Care Network (PCN)). Service leads consulted local voluntary sector services, schools and communities (See Supplementary File 1 for summary of the findings of the community consultation). Ultimately a localised clinic was developed that combines a collocated GP, paediatrician, and early years (Early Help) family support worker that CYP access following referral from their usual GP. The service is summarised in Figure 1 (and the supporting logic model can be found in Supplementary File 2). The three key components being delivered are preventative health advice, clinical consultations, and Early Help social support and the key components of each of these service offers is summarised in Box 1.

**Figure 1:**
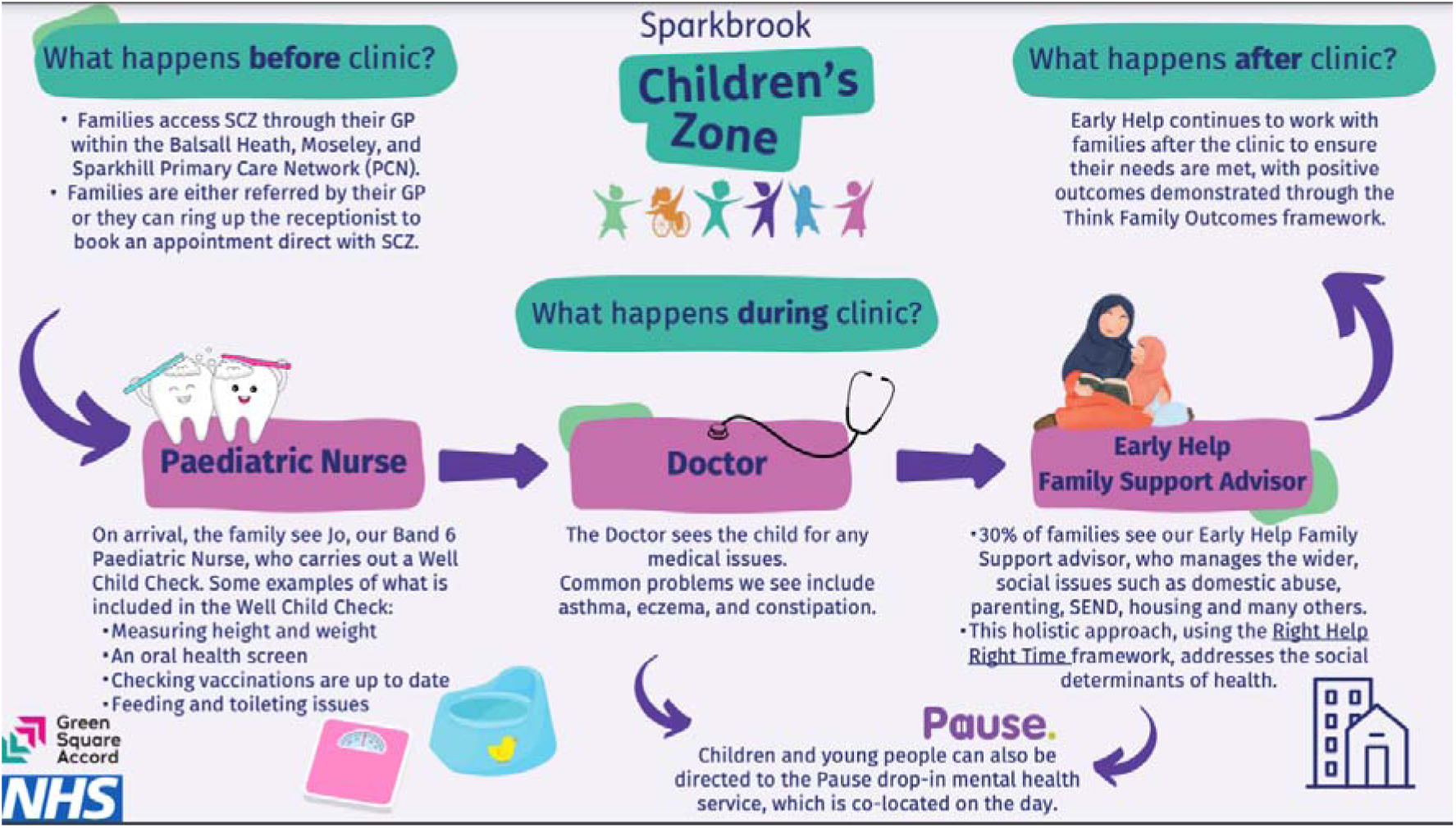
Sparkbrook Children’s Zone clinic model

### Staffing the service

The clinic comprises collocated paediatricians, GPs, and early years (Early Help) family support workers. Two paediatricians were provided by Birmingham Children’s Hospital each able to offer a half-day a week to manage and run clinics. The initial aim was to work alongside GPs from the partner PCN but after a few sessions, it became clear that they had limited capacity to commit to the clinic and so SCZ instead employed GPs (with an interest in paediatrics). The family support worker was provided by GreenSquareAccord. In February 2024 a paediatric nurse (Band 6) joined the team to help improve the preventive health offer embedded in clinics. She also worked alongside Early Help professionals to perform outreach work to bring the preventive health offer out to the community in schools, faith centres and other local settings. A drop-in mental health service for CYP (PAUSE [17]) attend on a weekly basis. They see any person registered with a Birmingham GP, who is under 25 years and is who is not experiencing a mental health crisis

#### Box 1

Summary of the key components of each element of the SCZ integrated care offer.

##### Preventive health care

- Smoking cessation advice (if there is a smoker in the household)
- Immunisation advice
- BMI calculated for CYP >2 years (and onward referral to a healthy eating/exercise programme when available/funded);
- An oral health check and a toothbrush pack (children who were amber/red on the screening check had support accessing a dentist if the child was not registered with one
- Healthy Start vitamins for CYP <4 years and mothers with an infant
- Directed via SMS post-clinic to the NHS Healthier Together website (https://www.what0-18.nhs.uk/)

##### Clinical appointments

- Clinic appointments are 25 minutes in length. GPs and paediatricians see all presentations except acute injuries (which are better managed in minor injury units or EDs).
- Clinicians are encouraged to provide written care plans via SMS text for all CYP attending with asthma, eczema or constipation (following guidance from the UK’s National Institute for Health and Care Excellence NICE).
- Any child or family with social support needs are referred directly to the team’s family support worker
- Electronic consultations were offered

##### Early Help

- Usually referred directly after the clinical appointment but can be referred directly to Early Help by GP, school.
- One-hour slots.
- Early intervention to prevent problems for families escalating.
- Early Help workers navigate issues for families with schools, health, the voluntary sector, sexual health services and many others to enable families to thrive.
- Early Help provides a broad range of services for CYP and their families including counselling and support for SEND; domestic violence; food poverty.,

### Data collection/analysis

There were approximately 14,000 CYP (i.e. <16 yrs old) registered to the PCN’s eight practices. A template embedded in the EMIS IT system was used to record patient encounters. Data was collected from March 2022 when the SCZ consisted of one weekly half-day clinic at a single site, expanded February 2023 to two half-day clinics a week and from October 2024, an additional two twilight clinics were added. Data from Early Help was collected on the team’s Empowering Communities with Integrated Network Systems (ECINS) IT system beginning April 2024.

Aggregate data on demographics (including ethnicity and deprivation score) and total number of appointments offered and attended were collected. Further to this, data was collected on the three key components of the SCZ service offer:

1. Preventative health - consisting of a range of advice and healthy start vitamins.
2. Clinical health care – specifically diagnoses/conditions, the provision of care plans for three common chronic conditions (asthma, constipation and eczema), alongside disposition, including referrals to secondary care, acute care, and Early Help
3. Social support – data were captured on the number of CYP referred, the reason for referral to Early Help and whether the family received light touch, intensive support or were escalated to Children’s Social Care.

Descriptive statistics were used to analyse aggregated and anonymised data collected by clinical and Early Help staff for all three arms of the SCZ offer – clinical health, preventive health, and social support.

### Patient and Public Involvement

Alongside ongoing community engagement (see Supplementary File 2), our mixed methods study has two patient and public involvement and experience representatives from Sparkbrook. They reviewed both the study protocol and have contributed to continued review of the study at meetings held in 2023 and 2024.

### Ethical approval

This study received UK Health Research Authority approval (Research Ethics Committee reference: 25/PR/0168).

## Results

The demographic profile of patients who attended SCZ clinics, the attendance/did-not-attend data, the numbers receiving preventive health, the number of CYP seen by clinical staff, family support and/or visited the drop in mental-health service.

### Patient demographics and attendance rates

Of the 2,265 CYP who attended the SCZ between March 2022 and December 2024: 1,042 (46%) were female; 314 (13.9%) were aged <12 months, 763 (33.7%) 12-59 months and 1,332 (58.8%) >60 months. There were limited data on ethnicity but for the 312 patients where this was recorded, the population was highly diverse, the largest proportion of patients were Pakistani (178, 57%). The Index of Multiple Deprivation (IMD) scores available for the total cohort (n=1,710) were overwhelmingly in the most deprived quintile i.e., 1,527, (89%), 115 (6.7%) in quintile 2 and 80 (4.7%) from quintile 3.

From opening the clinic from March 2022 to end December 2024, SCZ offered 2,423 face-to-face (F2F) clinic slots, of which 2,265 were taken up (93.5%). Of the clinic slots booked, 333 (14.7%) families did not attend (DNA, see discussion below).

#### 1) Delivery of preventive health

Families received a number of health promotion interventions with the highest figures being BMI measured – 83%; Healthy Start vitamins (<4yrs) – 72%; oral health screening/promotion – 39.7% and immunisation advice – 29% (compared to the PCN average of 1%). Figure 2 plots the number of CYP who received a health promotion intervention against total clinic attendances for each quarter 2023-24, noting that the paediatric nurse now primarily responsible for delivering preventative care joined the clinic in Q4 2024.

**Figure 2:**
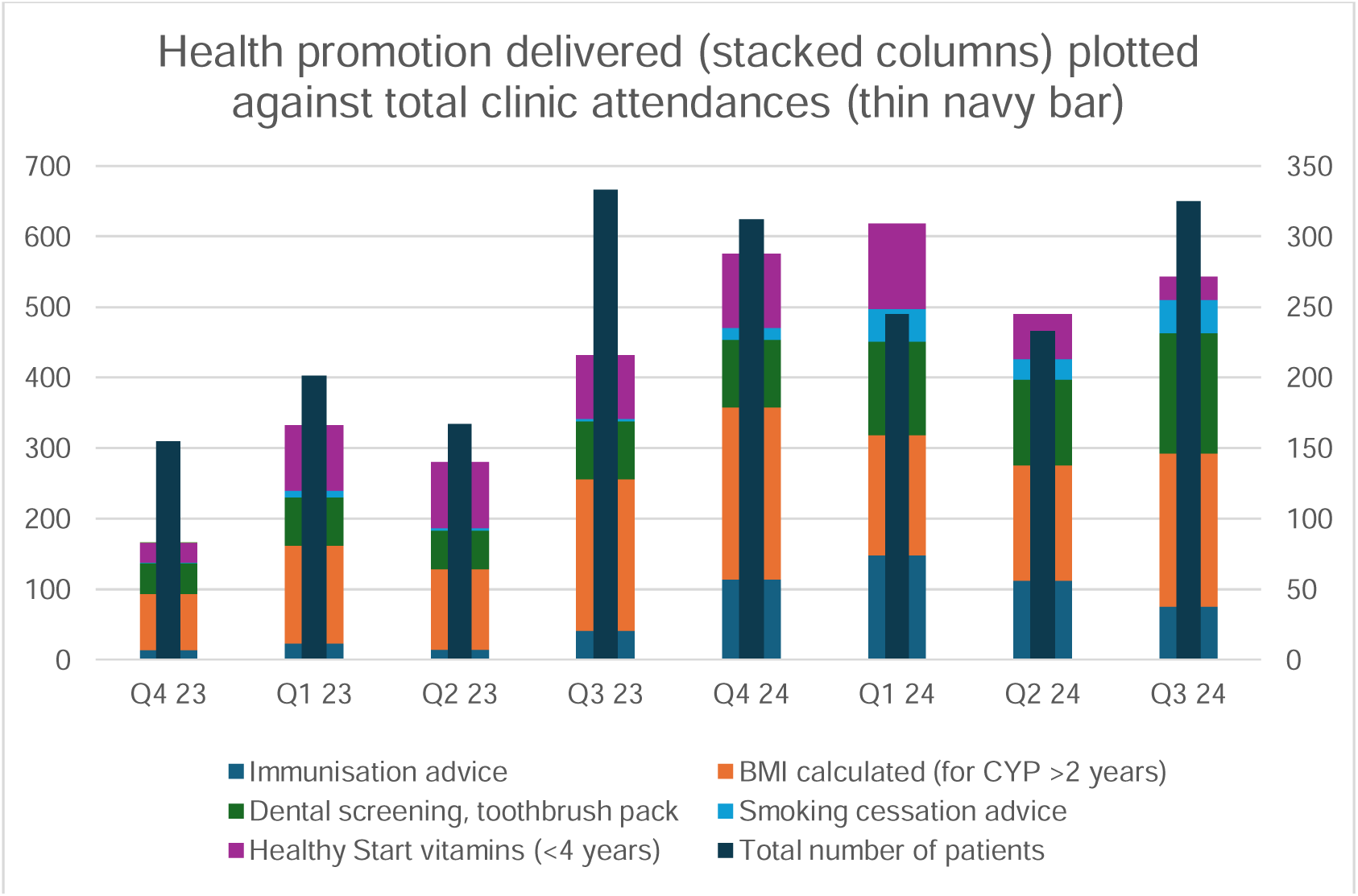
Health promotion work delivered by clinicians in F2F clinics.

#### 2). Clinical care

##### Reason for referral

The commonest issues referred to the SCZ clinic were behavioral issues; mental health concerns; growth and development; picky eating and obesity; asthma and acute respiratory infections; rashes, including eczema; autism and ADHD; speech delay; headache; abdominal pain, including constipation. For this period, we recorded 37 referrals to the collocated mental health drop-in service (PAUSE).

##### Provision of care plans

The SCZ clinic aimed to provide written care plans via SMS text for all CYP attending with asthma, eczema and/or constipation. Of the CYP attending with these conditions (March 2022 to December 2024), 91.5% of CYP with asthma received a care plan; 82% with eczema; 65.7% with constipation (see Figure 3).

**Figure 3:**
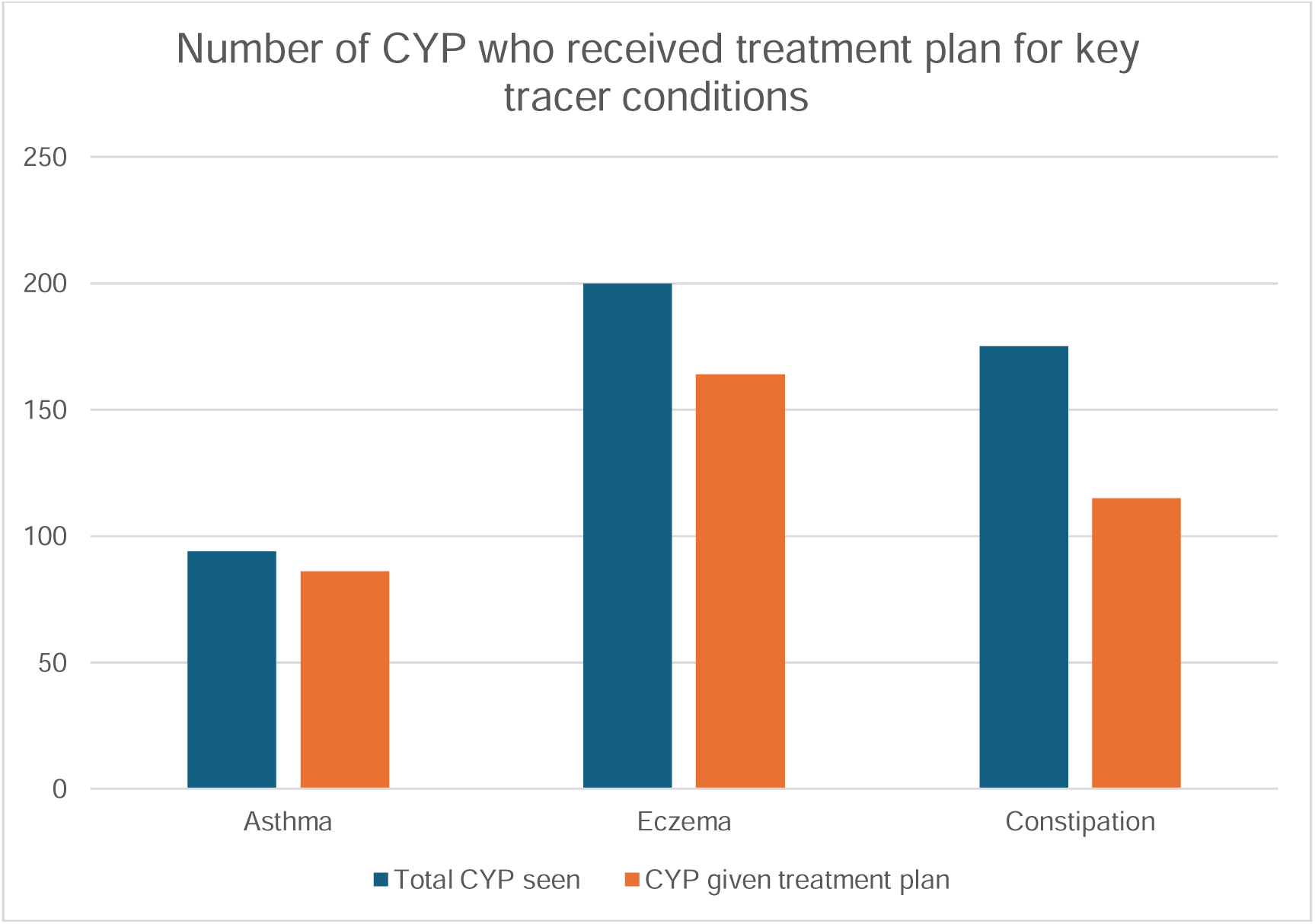
Number of CYP with asthma, eczema and/or constipation who received a written care plan via SMS text after F2F clinic.

##### Patient disposition from clinical consultations

Nearly three quarters of CYP who attended a F2F clinic (73.8%) were discharged back to their GP, with 6.4% brought back to an SCZ clinic (see Table 1), 3.8% of CYP were referred to secondary care and 1.1% required an emergency, same-day referral.

**Table 1:** Disposition outcomes for CYP attending Sparkbrook Children’s Zone F2F clinics, March 2022-December 2024

| Disposition | Total (out of 2,265 F2F attendances –includes DNAs, n=333) | % |
| --- | --- | --- |
| Discharged | 1,672 | 73.8% |
| SCZ follow-up | 146 | 6.4% |
| Referred to secondary care | 86 | 3.8% |
| Emergency hospital admission | 25 | 1.1% |
| Referred for Early Help support | 639 | 28.2% |
| Did not attend (DNA) | 333 | 14.7% |

There were 10 slots a week offered for electronic consultations but the take-up was lower than expected of and so this was discontinued after two years. Out of 390 slots taken, 149 (38%) of referrals were requests for an SCZ clinic appointment but only 33 (8.5%) resulted in a secondary care referral.

#### 3) Early Help

##### Reason for referrals

Of the CYP attending clinics, 639 (28.2%) were referred to the collocated family support worker (Early Help). Full data on outcomes was only available from April 2023 until March 2025 following the appointment of our programme manager. During this 24-month period, 548 children and their families were seen in clinic, of whom 490 (89%) were in the lowest IMD quintile; 302 (53%) were under 5 years of age; 173 (31%) were aged 6-11 years and 83 (15%) were aged 12-16 years.

The issues with which families most frequently presented were helping to find activities (e.g, stay-and-play groups, youth clubs) for CYP (134, 12%); behavioural issues (137, 12%); parenting skills (120, 11%); feeding issues (161, 15%); and special educational needs and disabilities (SEND) support (121, 11%). These are summarised in Figure 4.

**Figure 4:**
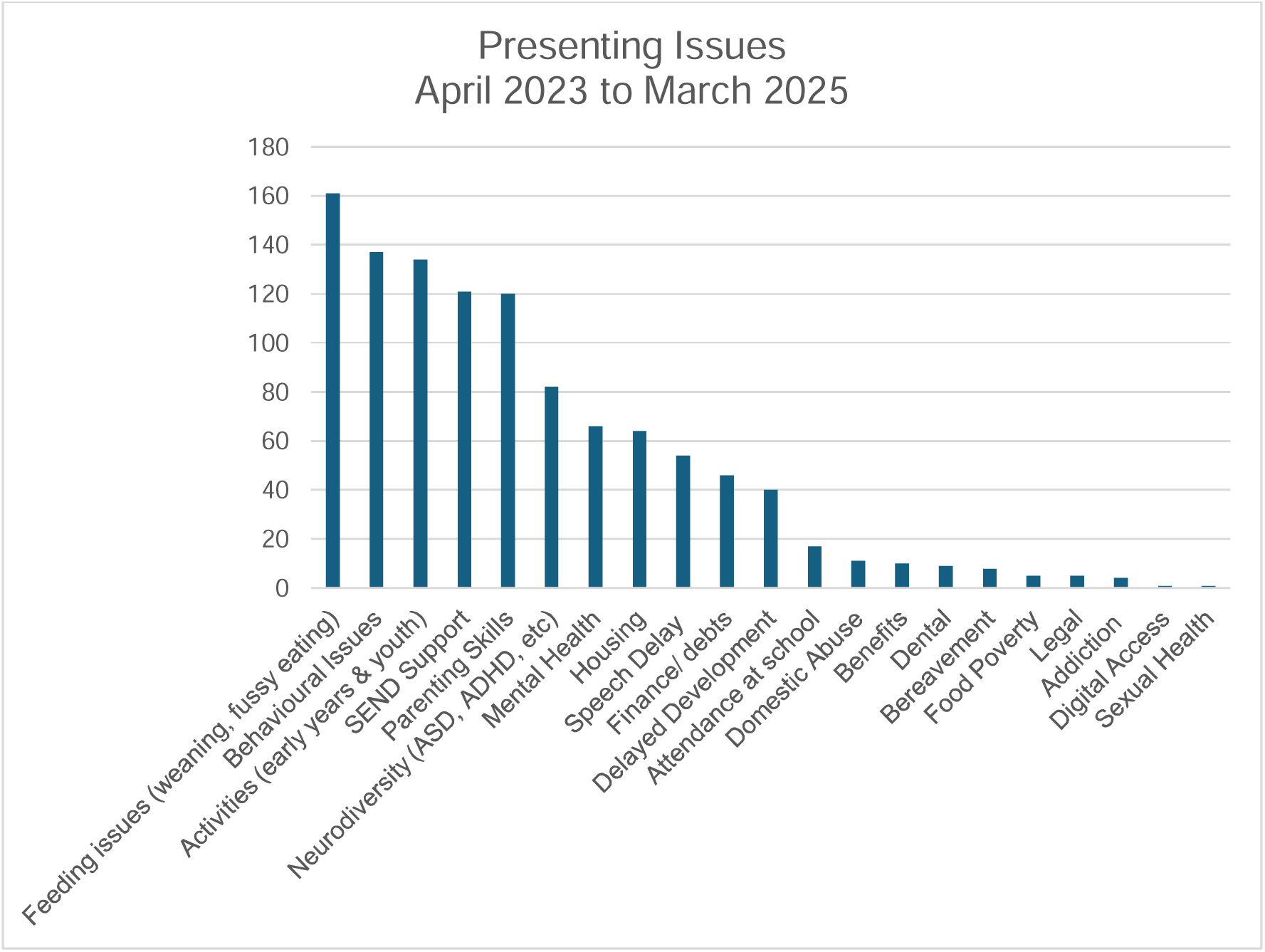
Issues presenting to SCZ’s family support worker in clinic (families can present with more than one issue), April 2023-March 2025

##### Referrals out of Early Help

Out of 548 of the total referred from April 2023-March 2025, 351 (64%) had “light touch” navigation to local support services and 197 (36%) had more intensive, multidisciplinary support (where a Family Connect Form was completed). Of the CYP requiring MDT support, one child was escalated to and managed by Children’s Social Services.

## Discussion

### Summary findings

The model of neighbourhood health for CYP demonstrated by the SCZ meets UK government and NHS aims for the provision of multi-disciplinarity neighbourhood health [10]. The SCZ experienced challenges in the practical delivery of the various elements of the service relating to the lack of inter-organisational infrastructure and reliable funding as described elsewhere [11]. Despite this, the SCZ pilot data has demonstrated significant take-up of preventative health, clinical health, and social support. Below we place our findings in the context of the existing evidence exploring the localised delivery of care for the underserved in each of the three service domains.

Of the total number of CYP who attended clinic, nearly three quarters were discharged, a figure close to that reported in a similar integrated care offer delivered in London (Connecting Children 4 Care (CC4C)[18]. We estimate that around 26% of clinic attendances were for acute medical conditions that could have been managed in routine primary care (although CYP benefited from the preventative and social support offers), and there was anecdotal evidence that partners in the PCN were filling consultation slots in the SCZ when their practice had exhausted capacity.

### Attendance

The most recent figures indicate that that some 15% of appointments are missed each year in general practice [19] with the majority of these missed due to patient factors with several studies in the UK showing a strong association between DNAs and minority ethnic populations, and lower socio-economic status [20]. The levels of DNA in the SCZ were close to this national average, though it’s worth observing that Sparkbrook has a super-diverse and socio-economically challenged patient population [21].

An increase in DNAs around school holidays was observed perhaps due to additional child care pressures as observed in other localities [20]. The SCZ data also linked DNAs to appointments for acute problems (e.g., a sore throat) which may have resolved prior to the appointment as also witnessed previously in primary care [22].

### Delivery of preventive health

In the UK health policy advocates healthcare professionals “make every contact count” by promoting preventive health in routine clinical consultations [23]. However, there are many barriers to delivering health prevention in standard primary care settings, including a lack of resource, adequate training, and lasting commitment from commissioners (funders) needed to support its implementation [24]. The SCZ was able to deliver key preventative health messaging and interventions for CYP and their families with this success attributable at least in part to the longer standard appointment time, which allowed greater opportunity to understand the patient and recognise the preventative care which was most relevant and appropriate [25].

The delivery of preventative care was also boosted by the recruitment of a paediatric nurse, with extensive neonatal and health visitor experience (as shown in Figure 4 from Q4, 2024: January-March). Their training and background meant they could confidently and independently advise on preventative health without recourse to a doctor or a family support worker. This reflects evidence elsewhere of the value of paediatric nurses in primary care environments in promoting healthy behaviours and offering a range of advice to families [26].

The reductions in vaccine uptake particularly in ethnic minorities, is a continuing and widely recognised public health issue in the UK and beyond [27]. The SCZ’s paediatric nurse increased the delivery of immunisation advice and the job role of paediatric nurse appears well placed to address parental hesitancy toward vaccination [28].

### Clinic delivery

The electronic consultation appeared to be poorly communicated and understood by GPs with uptake lower than anticipated. A number of barriers to the use of e-consultations by primary care staff have been described including concerns about increased workload, privacy, unsuitable infrastructure, and lack of clinical quality [29]. Though there are a number of known influences on GP referral rates including the characteristics of patients and GPs, the factors impacting teleconsultations on referral rates are not fully understood, nor is their impact on patient outcomes [30].

The lack of unsuitable referrals to the e-consultations by clinicians in other areas of the PCN raises an issue previously explored in other models of integrated care that not only does awareness need raising in patients but also amongst other parts of the health service [31]. This messaging to other care providers is also important in the context of evidence that suggests for new care offers to succeed they need to work hard at establishing understanding and trust in the surrounding service [32].

UK’s National Institute for Health and Care Excellence (NICE) recommends treatment plans to support more effective service utilisation and self-management [33] and the SCZ was able to deliver treatment plans to 91% of CYP with asthma. This compares favourably with the findings of a Global Asthma Network study which reported that these plans were typically provided for only 62.8% of patients [34].

Around one in five CYP in the UK is thought to have a mental disorder with impacts on individuals, their families, and for service utilisation [35]. To meet this need the SCZ embedded a weekly drop-in mental health service, which was greeted favourably by staff, and anecdotally by patients [11]. However, there were difficulties in precisely recording referral rates into the service with some being made by GPs and others by the SCZ’s family support workers. This highlights the challenges of sharing information across health and social care sectors which the SCZ shares with similar multi-agency care offers due to issues of governance and data interoperability [36, 37].

### Social support

Although there are multiple examples of integrated care for CYP in high-income countries few have adopted SCZ’s model of incorporating social support [6, 37]. The SCZ’s collocation of Family Support Workers, is one of the first examples in the UK of combining (and collocating) these services. While 28.2% of CYP who attended clinics were referred to a family support worker, it is feasible that the actual number of CYP impacted by the subsequent intervention might be triple that, as an intervention that might improve an aspect of their domestic circumstances would also benefit their siblings.

The collocation of health and social support has been shown to improve the financial security of families [9] and is cost effective for service providers [13]. However, a previous qualitative exploration of the SCZ reported difficulties in securing funding for the social support element [11], reflecting previous evidence of how funding restrictions, organisational barriers, and the undervaluing of social support in health, have hindered previous attempts of such integration[38].

#### Strengths and Limitations

The study is limited by the lack of a comparative figures from other models of integrated care because its novel approach to collocating social support has not been evaluated anywhere else in the UK [18]. There are also acknowledged data gaps around the collection of prospective data on referral into social support, or the take-up of the mental health support offer. Similarly, there were previously acknowledged gaps in the routinely collected data that prevented a more detailed breakdown of the conditions seen and the impact of the service on ED attendances [39].

## Conclusion

SCZ is a scalable intervention that integrates neighbourhood-based health services and social support. This descriptive study begins to build on a sparse evidence base around the successes and challenges of implementing an integrated multi-agency neighbourhood health service for CYP. The data is preliminary but understanding the nature of the shortcomings of the routinely collected data sets provides important learning for future evaluation of multi-disciplinary neighbourhood health and the study team is continuing to collect quantitative data on the performance of the SCZ and capture qualitative data on the experience of users.

### Authors statement

## Acknowledgments

The authors wish to thank the community in Sparkbrook for their continued engagement with and contributions to the Sparkbrook Children’s Zone.

## Ethics statement

Ethical approval was granted by Health Research Authority 25/PR/0168.

## Author Contributions

Chris Bird: Conceptualisation, Methodology, Investigation, Supervision, Data Curation, Formal analysis, Writing - original draft, Writing - review and editing

Frances Dutton: Investigation, Data curation, Formal analysis, Writing - review and editing

Simarjeet Kaur: Investigation, Data curation, Formal analysis, Writing - review and editing

Caroline Wolhuter: Investigation, Data curation, Formal analysis, Writing - review and editing

Ian Litchfield: Conceptualisation, Methodology, Writing - review and editing

## Conflict of Interest

The authors have confirmed no conflicts of interest

## Data Availability Statement

The datasets used and/or analysed during the current study are available on request from the first author, in line with the ethical approvals.

## Study Funding

The study was funded by Birmingham Women’s and Children’s Foundation NHS Trust

## Notes

### Competing Interest Statement

The authors have declared no competing interest.

### Funding Statement

The Study was funded by Birmingham Women's and Childrens NHS Foundation Trust

### Author Declarations

This study received UK Health Research Authority approval (Harrow Research Ethics Committee reference: 25/PR/0168).

