## Supplementary figures and images for "Early activity and impact of a neighbourhood multidisciplinary team that integrates health, and social support for underserved children and young people in Birmingham, UK: an observational study"

### Supplementary File 1

**Supplementary File 1: Logic model for SCZ intervention**

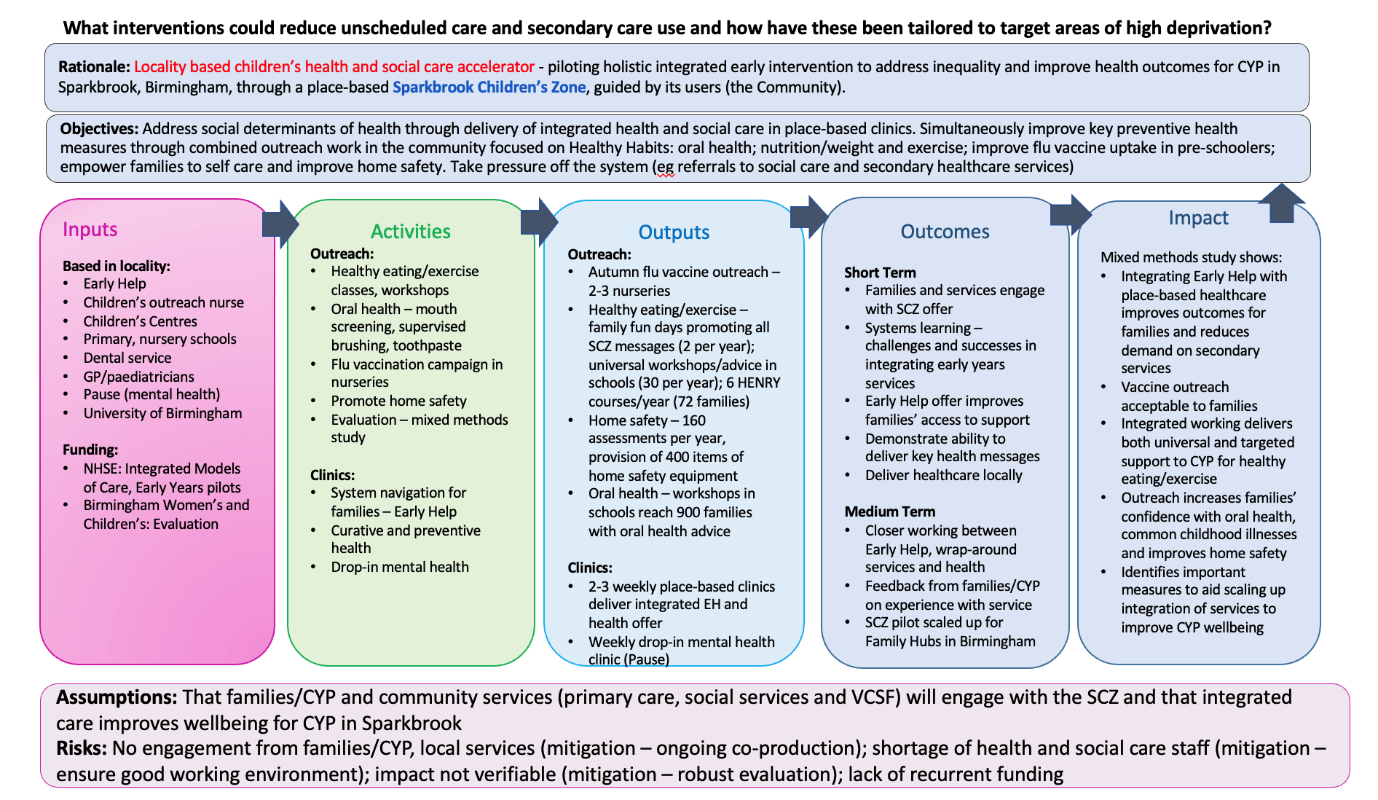
