## Supplementary File 2 for "Early activity and impact of a neighbourhood multidisciplinary team that integrates health, and social support for underserved children and young people in Birmingham, UK: an observational study"

**Supplementary File 2: Main issues raised around health and social support during community engagement exercise in Sparkbrook in 2021**

Medical Support Needs

|  |  |
| --- | --- |
| Get medicine / prescription | 11 |
| Depression / anxiety | 10 |
| Asthma / breathing | 7 |
| Ecsema / skin problem | 7 |
| Baby check | 4 |
| Coughs | 4 |
| Get medical advice | 4 |
| Stomach ache | 4 |
| Tooth ache | 4 |
| High temperature | 3 |
| Tooth ache | 3 |
| Vacination / Immunisation | 3 |
| Weight management | 2 |
| Weight Management | 2 |
| Constipation | 1 |
| Drugs and alcohol | 1 |
| Minor injuries | 1 |

**Figure A Medical support needs of local community**

Other Support Needs

|  |  |
| --- | --- |
| Mental Health | 11 |
| Skills & training | 10 |
| Housing | 8 |
| Education | 7 |
| Data & IT | 6 |
| Domestic Abuse | 6 |
| Parenting | 6 |
| Food & Energy | 5 |
| Sexual Health | 5 |
| Under 5's Support | 5 |
| Uniform & Clothes | 5 |
| Employment support | 4 |
| Youth Support | 4 |
| Play | 3 |
| Special Needs | 3 |
| Money & debt advice | 2 |
| Volunteering | 2 |

**Figure B Non-clinical support needs of local community**
